# Comparative analysis of human immune responses following SARS-CoV-2 vaccination with BNT162b2, mRNA-1273, or Ad26.COV2.S

**DOI:** 10.1101/2021.09.21.21262927

**Authors:** Dominique J. Barbeau, Judith M. Martin, Emily Carney, Emily Dougherty, Joshua D. Doyle, Terence S. Dermody, Alejandro Hoberman, John V. Williams, Marian G. Michaels, John F. Alcorn, W. Paul Duprex, Anita K. McElroy

## Abstract

**Background:** Three SARS-CoV-2 vaccines, two based on mRNA, BNT162b2 and mRNA-1273, and one based on an adenovirus platform, Ad26.COV2.S, received emergency use authorization by the U.S. Food and Drug Administration in 2020/2021. These vaccines displayed clinical efficacy in initial studies against confirmed COVID-19 of 95.0%, 94.1%, and 66.9%, respectively.

**Methods:** Individuals receiving one of these vaccines were invited to participate in a prospective longitudinal comparative study of immune responses elicited by the three vaccines. In this observational cohort study, humoral responses were evaluated using a SARS-CoV-2 receptor-binding domain (RBD) ELISA and a SARS-CoV-2 virus neutralization assay at mean of 21-31 days and 45-63 days following each initial vaccination. IFN-γ ELISPOT assays were conducted with peripheral blood mononuclear cells obtained at a median of 45-63 days after each initial vaccination.

**Results:** The two mRNA-based platforms elicited similar RBD ELISA responses and neutralizing antibody responses. The adenovirus-based vaccine elicited significantly lower RBD ELISA and SARS-CoV-2 virus neutralization activity. The mRNA-1273 vaccine elicited significantly higher spike glycoprotein-specific T cell responses than either the BNT162b2 or the Ad26.COV2.S vaccines.

**Conclusions:** Both mRNA based vaccines elicited higher magnitude humoral responses than Ad26.COV2.S and mRNA1273 elicited the highest magnitude of T cell response. Neutralizing antibody titers correlated with reported estimates of vaccine efficacy.

**Summary of key points:** We compared antigen specific humoral and T cell responses following vaccination with BNT162b2, mRNA-1273, or Ad26.COV2.S. Both mRNA based vaccines elicited higher magnitude humoral responses than Ad26.COV2.S and mRNA1273 elicited the highest magnitude of T cell response.

## Introduction

Two mRNA-based vaccines (BNT162b2 and mRNA-1273) and one adenovirus-based vaccine (Ad26.COV2.S) have been used in the US since EUA was granted for each (December 2020 for the mRNA vaccines and February 2021 for the adenovirus vaccine). While data have been published for each of these vaccines confirming safety, immunogenicity, and efficacy [1-7], only limited data are available that compare vaccine-induced immune responses amongst the three vaccines using identical immunological assays [8]. In this study, we assessed a cohort of SARS-CoV-2 naive individuals who received BNT162b2, mRNA-1273, or Ad26.COV2.S vaccines for antigen-specific humoral and T cell immunity using identical immunologic assays to allow direct comparisons of the elicited responses. To date, an immune correlate of protection has not been established, however, neutralizing antibody titers may be a possible correlate [9]. Mathematical modelling combined with analysis of published immunogenicity data has been used to demonstrate that the ratio of neutralization titer following vaccination to that during convalescence following naturally-acquired disease correlates with the reported clinical efficacy for each vaccine [10]. Our study demonstrates a correlation between neutralizing antibody titer and reported vaccine efficacy, but we did not observe this correlation with IFN-γ ELISPOT responses, supporting the use of neutralizing antibody titer as a correlate of protection.

## Methods

### Human subjects research

A convenience sample of participants 18 years of age and older were prospectively enrolled if they were planning to receive two doses of mRNA-1273 or BNT162b2, or a single dose of Ad26.COV2.S under FDA EUA between December 2020 and March 2021 either in an occupational (BNT162b2) or a community (mRNA-1273 and Ad26.COV2.S) setting. After obtaining written informed consent, whole blood was obtained in cell preparation tubes (BD) on the day of enrollment/first vaccination, and during follow up visits. Plasma and peripheral blood mononuclear cells (PBMCs) were isolated and cryopreserved using standard methods. Institutional Review Board approval was provided by the University of Pittsburgh Human Research Protection Office.

### Enzyme-linked immunosorbent assay (ELISA) and focus reduction neutralization assays (FRNT)

ELISA and FRNT assays were conducted as previously described [11]. Samples with FRNT_50_ titers below the limit of detection (LOD=10) were assigned a value of 5 for graphical depiction and statistical analysis on the figures.

### ELISPOT assays

PMBCs were incubated for 24 hours with 2 μg/mL of a SARS-CoV-2 complete spike glycoprotein mega pool consisting of 15-mer peptides overlapping by 11 residues based upon the original Wuhan-Hu-1 strain (GenBank MN908947.3) (Miltenyi) with 1 x Cell Activation Cocktail (without Brefeldin A, BioLegend) or media alone. Human IFN-γ ELISPOT assays were conducted according to the manufacturer’s instructions (Mabtech).

### Data analysis

Graphpad Prism was used for statistical analysis and to prepare figures. Four samples were missing from visit 2 for the mRNA-1273 participants. Samples were available for all other participants at all time points. To analyze ELISA data a mixed-effects model with the Geisser-Greenhouse correction and Tukey’s multiple comparison test was used. To analyze FRNT data, a mixed-effects model and Sidak’s multiple comparison test was used. A Kruskal-Wallis test with Dunn’s multiple comparisons test was used to compare ELISPOT data between vaccine groups. Sex, age, and racial/ethnic distributions of participants were compared with those of the catchment area using one-way chi-squared goodness-of-fit tests.

## Results

Participant age, sex, and race were provided at the time of enrollment (Table 1). Information about co-morbidities was collected from the medical record or directly from study participants.

**Table 1:**
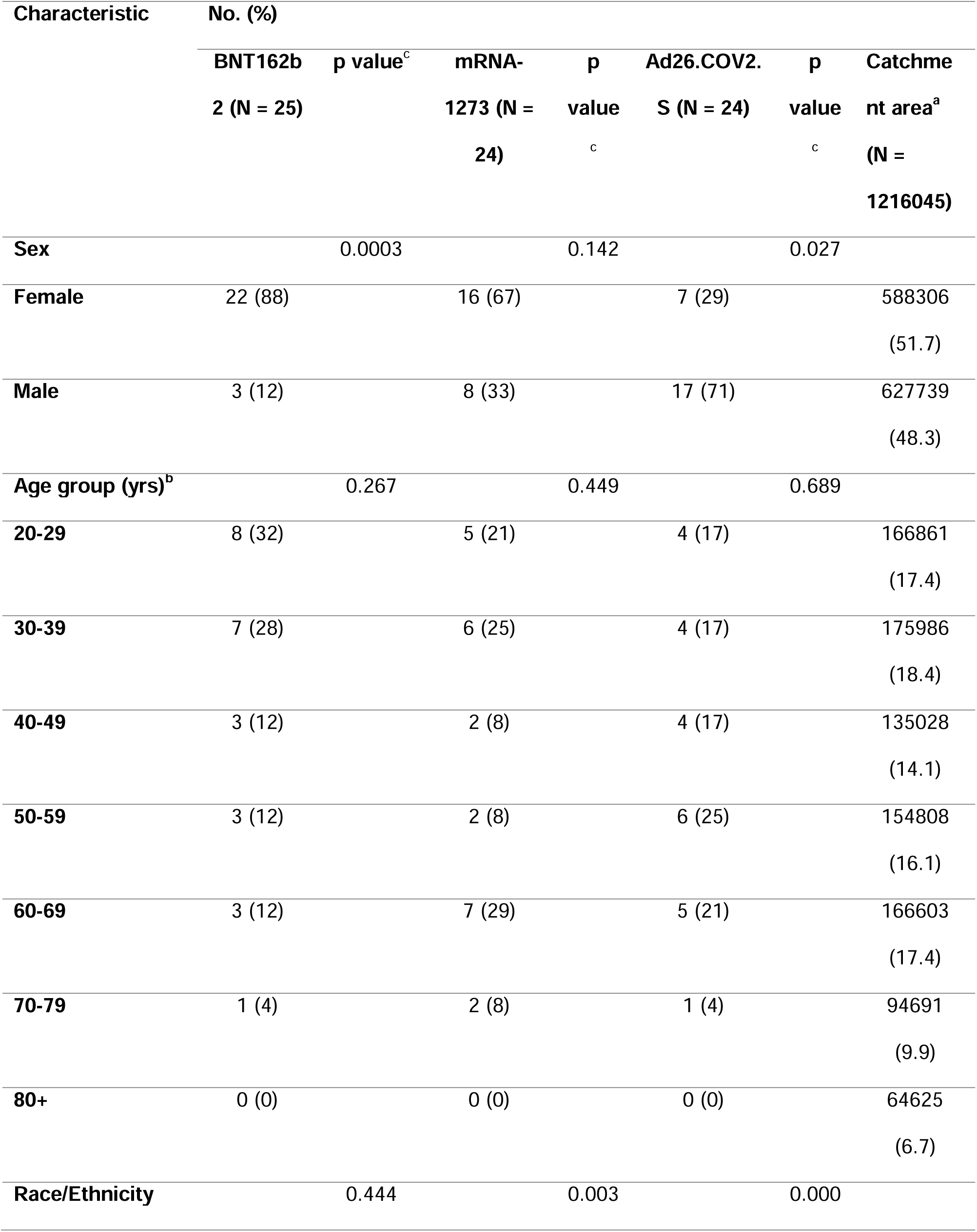
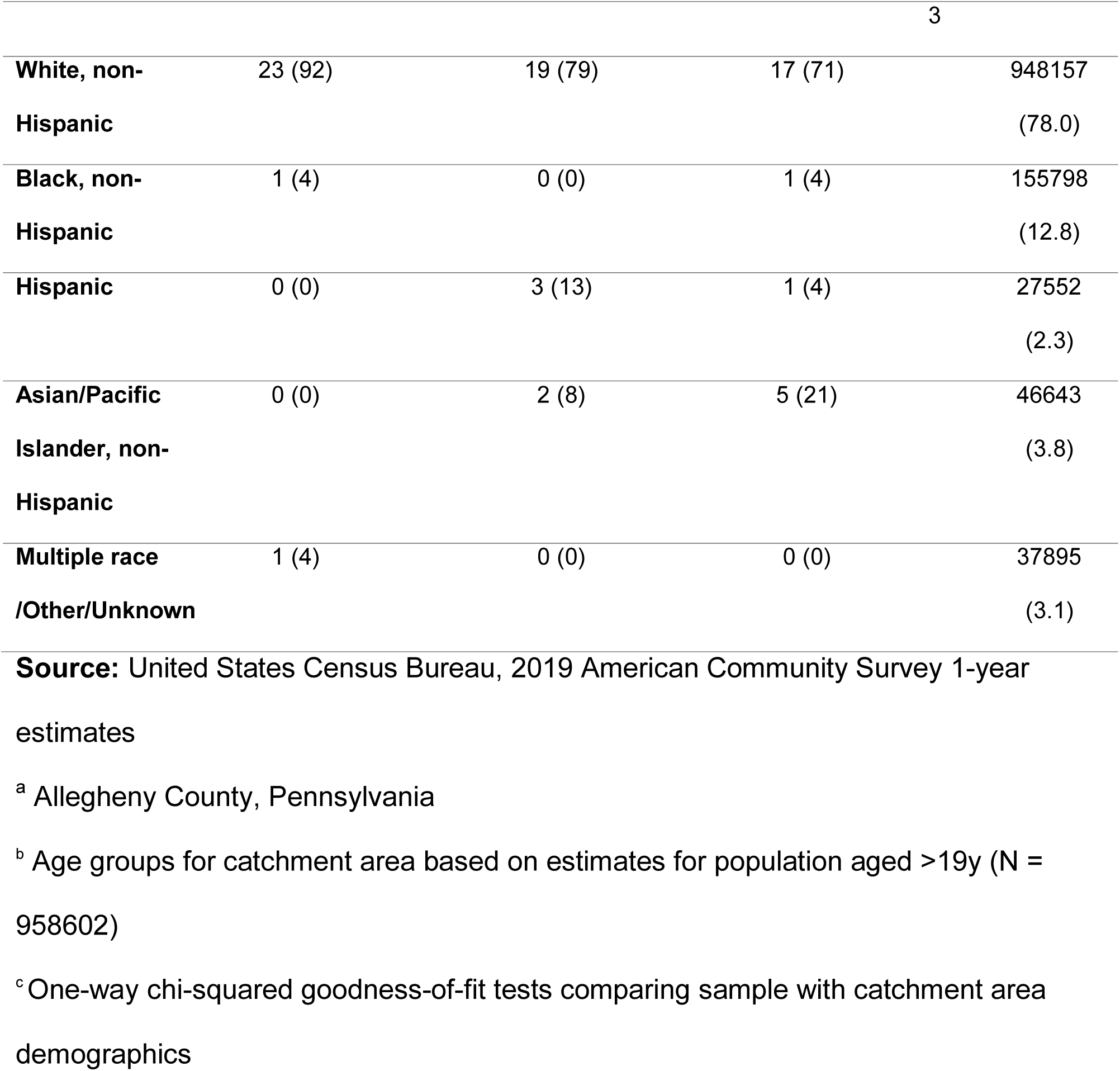
Demographics of the study population.

An RBD ELISA was conducted using plasma samples from all participants at all three time points (enrollment/date of first vaccination and at subsequent visits, Table 2). Participants with higher-than-average baseline RBD titers were tested by SARS-CoV-2 nucleoprotein (N) ELISA, and 5 individuals were found to have N-specific ELISA titers greater than or equal to 900 [11]. As this finding is consistent with prior SARS-CoV-2 infection, data from these participants were excluded from further analysis. One mRNA-1273 vaccine recipient relocated after enrollment precluding collection of additional data. After these exclusions, 25 BNT162b2 recipients, 24 mRNA-1273 recipients, and 24 Ad26.COV2.S recipients were included in the study. On the date of first vaccination (pre-bleed), 6 individuals had detectable RBD ELISA titers relative to a pre-pandemic normal human control group. There were no statistically significant differences in baseline titers between the vaccine groups (Figure 1). At visit 2 (median 21-31 days post initial vaccination), most study participants had notable antibody titers with the exception of two BNT162b2 recipients, one of whom was immunocompromised secondary to ongoing therapy for breast cancer with capecitabine. At visit 2, RBD ELISA titers were significantly higher in mRNA-1273 recipients compared with Ad26.COV2.S recipients (p=0.0008) At visit 3 (median 45-63 days post initial vaccination), following booster doses for both of the mRNA-based vaccines, the BNT162b2 titers were equivalent to the mRNA-1273 titers, and both were significantly higher than those achieved by the Ad26.COV2.S (single-dose regimen) (p=0.0059 for BNT162b2 vs. Ad26.COV2.S and p=0.0315 for mRNA-1273 vs. Ad26.COV2.S). RBD ELISA titers observed in these cohorts correlated with those previously reported in immunogenicity studies of the various vaccines with Ad26.COV2.S having 2-3 log GMT [6], and mRNA-1273 and BNT162b2 having 4-5 log GMT after completion of the two dose series [3, 12].

**Table 2:** Intervals between vaccination and follow up visits for each vaccine group.

| **Median # Days (Interquartile Range)** | **mRNA-1273** | **BNT162b2** | **Ad26-COV2.S** |
| --- | --- | --- | --- |
| **Interval between vaccine 1 and visit 2** | 28 (27-29) | 21 (17-22) | 31 (28-35) |
| **Interval between vaccine 1 and visit 3** | 63 (60-68) | 45 (43-46) | 60 (56-66) |
| **Interval between vaccine 2 and visit 3** | 36 (32-41) | 24 (22-25) | N/A |

**Figure 1:**
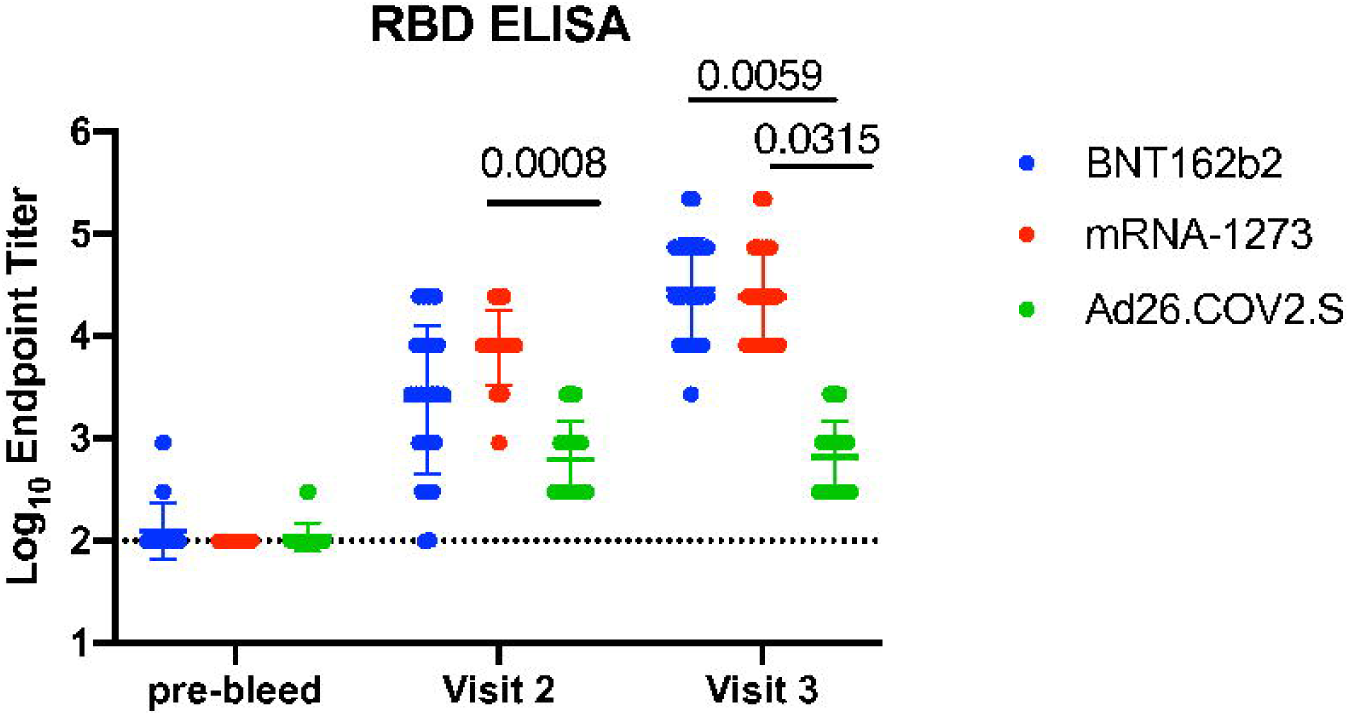
RBD ELISA titers amongst vaccine recipients. Plasma from each timepoint was tested by an RBD ELISA. Each data point is shown, the geometric mean and geometric standard deviation are plotted. The dotted line is the limit of detection of the assay. Statistically significant differences are noted by the respective p value.

Plasma samples from visit 2 and visit 3 were tested in a neutralization assay using SARS-CoV-2. FRNT_50_ titers were at or below the limit of detection (LOD=10) for a small subset of participants after a single immunization and did not differ significantly between vaccines (Figure 2A). Following a second dose of either of the mRNA vaccines, FRNT_50_ titers were significantly higher than those achieved with single dose Ad26.COV2.S vaccination (p<0.0001 for both BNT162b2 and mRNA-1273 vs. Ad26.COV2.S). A trend towards higher mean FRNT_50_ titer was observed following the vaccination series with mRNA-1273 than with BNT162b2, but this was not significant. Given the relatively low neutralization titers observed in some subjects, several observations did not meet the 50% focus-reduction threshold. Therefore, the data were analyzed as a function of percent neutralization of input virus at a constant 1:20 dilution of plasma (Figure 2B). This strategy allowed a more nuanced assessment of the capacity of individual plasma specimens to neutralize virus. Participants who received a single dose of mRNA-1273 achieved higher percent neutralization at 1:20 dilution than those receiving Ad26.COV2.S (p<.0001). Following the booster dose, BNT162b2 and mRNA-1273 vaccination elicited similar neutralizing capacity, and percent neutralization at 1:20 dilution elicited by both mRNA vaccines was higher in magnitude than that elicited by the single dose Ad26.COV2.S at visit 3 (p<0.0001 for both BNT162b2 and mRNA-1273 versus Ad26.COV2.S). Interpretation of these data in the context of previous independently published reports of each vaccine should take into consideration the differences in methods used in these studies. In some studies, pseudovirion assays are used, in others live virus is used but the readouts vary based upon the assay-luciferase reporters in some, foci in others. Additionally, if the inoculum (virus combined with serum) is left in place following adsorption, this could alter the assay to assess not only antibodies that block binding but also those that block cell to cell spread or post-binding steps in fusion as has been shown for antibodies that target the spike N-terminal domain [13, 14]. Slightly lower FRNT_50_ titers noted in our study could be because we remove the inoculum after a 1-hour incubation period, hence our FRNT assay is specific for antibodies that block binding to the cellular ACE2 receptor. For this reason it is important to have a comparator control group that can be used in all studies to permit normalizing between assays as has been proposed by WHO [15].

**Figure 2:**
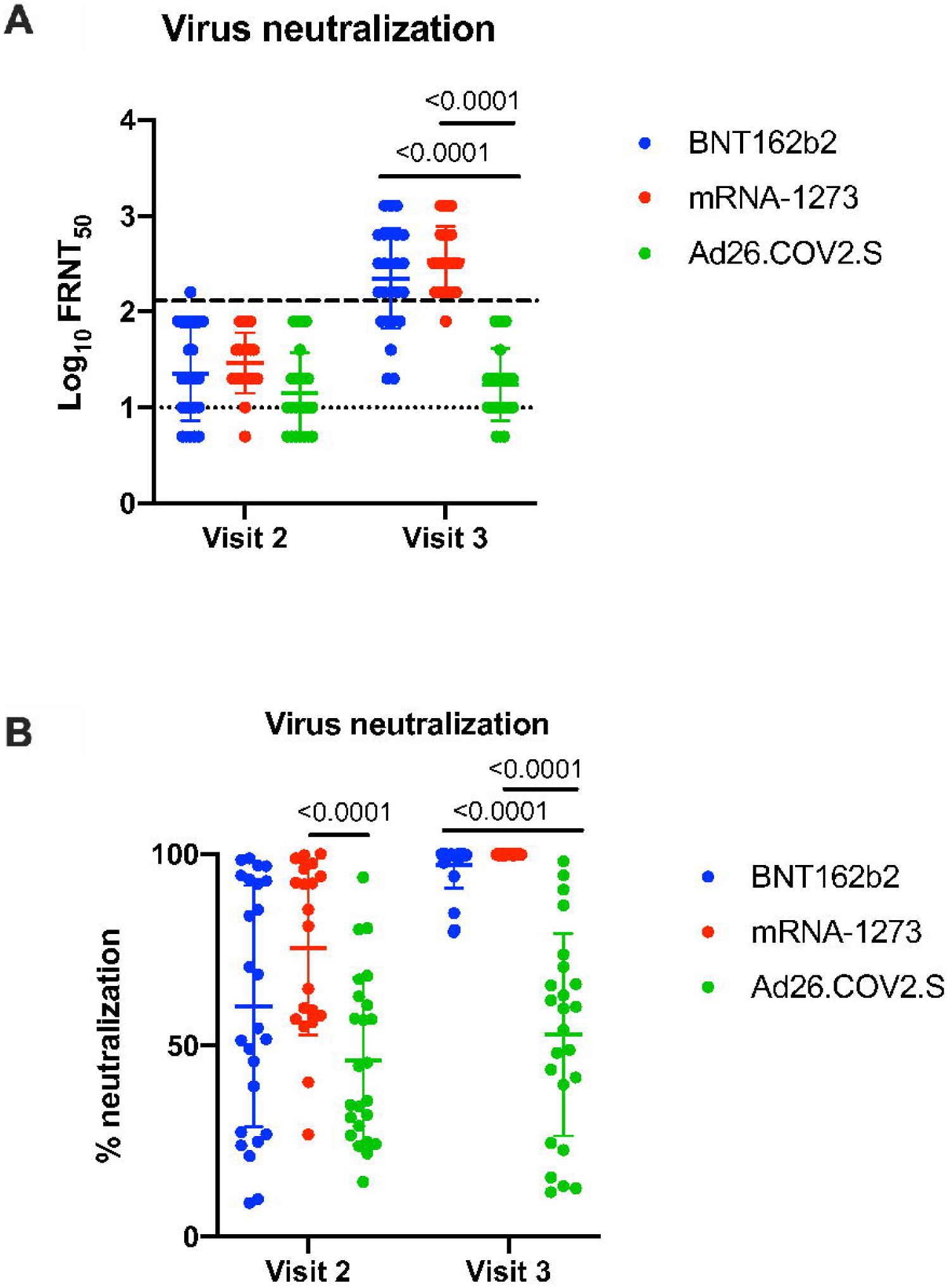
SARS-CoV-2 virus neutralization amongst vaccine recipients. Plasma from each timepoint was tested by a SARS-CoV-2 neutralization assay. Data are shown as FRNT_50_ (A) and as the percent of neutralization of input virus achieved at a 1:20 dilution of plasma (B). Each data point is shown, the geometric mean and geometric standard deviation (A) or mean and standard deviation (B) are plotted. Statistically significant differences are noted by the respective p value. The limit of detection for the FRNT_50_ assay is a titer of 10, indicated by a dotted line. The geometric mean titer (GMT) of a series of convalescent patients is indicated by a dashed line.

SARS-CoV-2 spike glycoprotein-specific T cell responses were assessed using an IFN-γ ELISPOT assay with PMBCs from study participants at visit 3 (Figure 3). mRNA-1273 recipients had a significantly higher magnitude of IFN-γ producing T cells following *ex vivo* stimulation of PBMCs with a total spike peptide mega pool when compared with BNT162b2 and Ad26.COV2.S recipients (p<0.0001 for BNT162b2 vs. mRNA-1273 and p=0.0001 for Ad26.COV2.S vs. mRNA-1273). No significant differences in T cell responses were apparent between the BNT162b2 and Ad26.COV2.S recipients.

**Figure 3:**
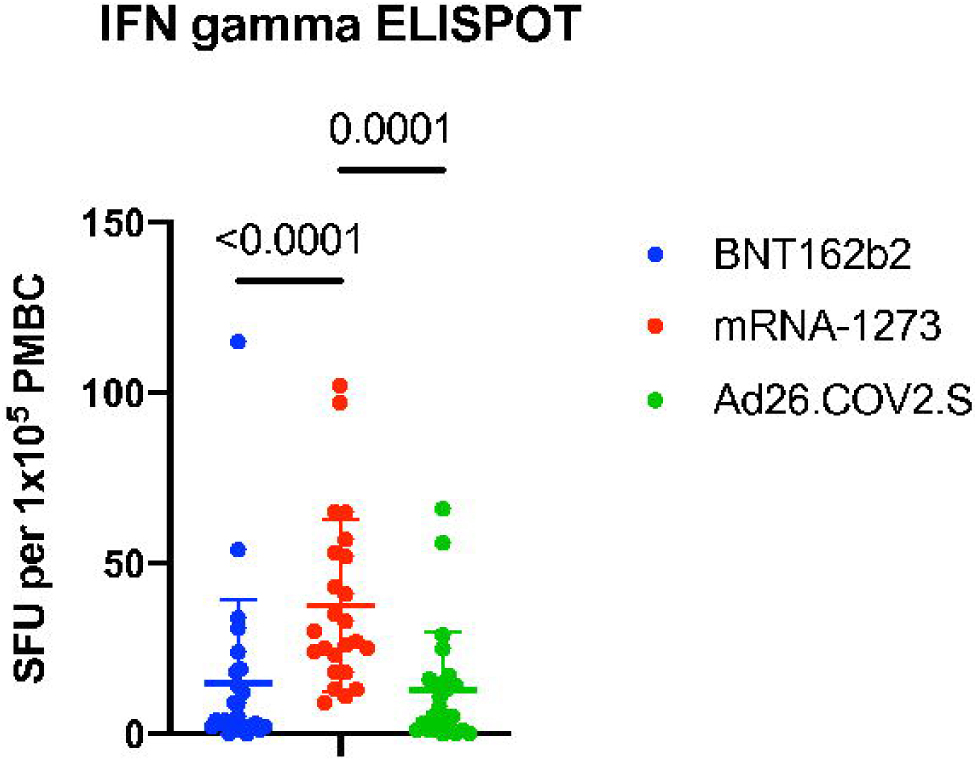
SARS-CoV-2 spike protein-specific T cell responses amongst vaccine recipients. PBMCs from the visit 3 timepoint were tested by an IFN gamma ELISPOT assay. Data are shown as spot forming units (SFU) per 100,000 PBMCs. Each data point is shown, the mean and standard deviation are plotted. Statistically significant differences are noted by the respective p value.

## Discussion

### Comparative humoral immune responses correlated with published efficacy data

The mRNA-1273 vaccine trended towards the greatest magnitude of spike-specific humoral immunity. This was also recently reported in another study of humoral immunity following vaccination [16]. The mRNA-1273 vaccine contains 100 μg of mRNA encoding the full-length, stabilized spike glycoprotein[2]. In our small cohort, BNT162b2 achieved a similar magnitude of antibody response after the second dose. The BNT162b2 vaccine contains 30 μg of mRNA encoding the full-length, stabilized spike glycoprotein [4]. The Ad26.COV2.S vaccine is administered at a dose of 5 × 10^10^ virus particles [5]. The adenovirus vector is replication-incompetent, thus, similar to the mRNA vaccines, only cells that take up the vaccine following inoculation will produce the SARS-CoV-2 spike glycoprotein. There is no amplification of these three vaccines *in vivo*. Data on humoral immune responses suggest a correlation between the dose of antigen and the kinetics and magnitude of the response, at least for the mRNA vaccines. Accordingly, a dose-response was reported in early Phase I/II clinical trials of all three of the vaccine platforms [3, 6, 7, 12]. However, as with all vaccines, the goal is to achieve a balance between reactogenicity and immunogenicity. Significant reactogenicity was reported in mRNA-1273 clinical trials and greater than 50% of study subjects reported a systemic adverse reaction of any grade after the first dose and over 75% after the second dose [2]. The BNT162b2 vaccine also had significant reactogenicity [4] but with lower frequency than mRNA-1273. Following single dose Ad26.COV2.S vaccination, over 60% of participants reported a systemic adverse reaction of any grade [5]. Significant reactogenicity in the setting of lower immunogenicity with the adenovirus vaccine might be attributable to pre-existing immunity to adenovirus or activation of innate immune pathways that differ from the mRNA vaccines.

A ratio of vaccine/convalescent neutralization titer may represent an immunologic surrogate of protection [10]. In our previous work evaluating neutralization titers of individuals with serologically-identified prior infection (n=30), we found a FRNT_50_ GMT of 130 [11]. Similarly, we measured a FRNT_50_ GMT of 129.6 and in a cohort of virologically confirmed outpatients (n=56) (unpublished data). The FRNT_50_ GMT for each vaccine at visit 3 for samples with a positive value was 223.2 for BNT162b2, 348.9 for mRNA-1273, and 20.7 for Ad26.COV2.S. These values would result in a ratio of vaccine/convalescent titer of approximately 1.7 for BNT162b2, 2.7 for mRNA-1273, and 0.2 for Ad26.COV2.S, which would equate to a predicted protective efficacy of >90% for the two mRNA vaccines and approximately 50% for the adenovirus-based vaccine as extrapolated from Khoury et al [10]. These predicted efficacy estimates are similar to the observed estimates of 94.1-95.0% vaccine effectiveness for the two mRNA vaccines [2, 4], providing support for the use of vaccine/convalescent titer as a correlate of protection. However, our calculated value of predicted efficacy for the Ad26.COV2.S vaccine group (approximately 50%) was slightly lower than that observed in the Phase III clinical trial for Ad26.COV2.S (66.9%) [5], which may be attributable to the fact that our Ad26.COV2.S cohort is mostly older men or it could be attributable to the difficulty of comparing data using different immunological assays and the lack of a clear definition of “convalescent” in the calculation of GMT from convalescent patients. Further studies are necessary to confirm a precise serum neutralizing titer that correlates with protection and to identify and standardize assays for this determination.

### Antigen-specific T cells do not necessarily correlate with vaccine efficacy

When our T cell data are compared with published data from initial immunogenicity studies for Ad26.COV2.S, the data are concordant with Ad26.COV2.S Phase 1-2a data showing IFN-γ ELISPOT mean of approximately 180 spot forming units (SFU) per 1×10^6^ PMBC [6] and our data showing a mean of 14 spot forming units per 1× 10^5^ PBMC. The initial immunogenicity trials for mRNA-1273 and BNT162b2 did not report on T cell studies. Since their publication, several studies have assessed T cells in older adults, immunocompromised hosts or in mixed cohorts of mRNA vaccine recipients and T cell responses; one study largely composed of BNT126b recipients had SFU ranging from the LOD to 1000 per 1×10^6^ PBMC [17] which is consistent with our data showing SFU ranging from 0 to 100 per 1× 10^5^ PBMC. This assay is another area in which standardization is needed for cross study comparison.

The finding of significantly higher levels of spike-specific T cells following vaccination with mRNA-1273 than with either BNT162b2 or Ad26.COV2.S could be attributable to higher levels of antigen or to the dosing regimen. A notable feature of all three vaccines is the intracellular expression of viral antigen, which allows intracellular processing and presentation of spike glycoprotein-derived peptides in the context of MHC I and stimulation of virus-specific T cell responses. Intracellular antigen expression is an advantage of viral-vectored vaccine platforms such as adenovirus-based vaccines. However, as our data demonstrate, mRNA vaccines elicit equivalent (BNT162b2) or higher-magnitude (mRNA-1273) antigen-specific T cell responses compared to Ad26.COV2.S. These data have an additional important implication. Clinical trials indicated that BNT162b2 has an efficacy of 95% after the two-dose series, while Ad26.COV2.S efficacy was 66.9%. Since these two vaccines elicit similar T cell responses in IFN-γ ELISPOT assays, these data suggest that the T cell component of the immune response is probably not responsible for the enhanced protection afforded by BNT162b2 versus Ad26.COV2.S. However, the IFN-γ ELISPOT assay did not discriminate between CD4+ and CD8+ T cells, and thus, we cannot exclude the possibility that a specific T cell subset or function plays a role in protection, or that T cell mediated protection plays a role in the context of reduced humoral immunity.

### Could antigen-specific T cells provide a protective advantage against variants of concern?

One of the most pressing problems in the pandemic is the emergence of viral variants of concern (VOC). Serum from vaccine recipients appears to neutralize the variants reported thus far, despite a reduction in viral neutralizing capacity in some cases [18-21]. Additionally, vaccinated individuals appear to be protected from severe disease and hospitalization following infection with VOC [22]. Studies of influenza vaccination suggest that heterologous protection is provided by virus-specific T cell immunity [23]. Most SARS-CoV-2 T cell epitopes are largely conserved between the variants [24], and even unexposed individuals have some virus-specific T cells due to cross-reactivity with epitopes conserved among common human coronaviruses [25]. Given the existence of virus-specific, cross-reacting T cells, we hypothesize that increased protection from disease caused by the variants could be provided by vaccines that also stimulate strong T cell responses in addition to strong humoral responses. If this were the case, mRNA-1273 could potentially provide better protection from variants than either BNT162b2 or Ad26.COV2.S. Further research will be required to test this hypothesis.

Limitations of our study include that it was an observational cohort study without randomization, and participants receiving the various vaccines were not matched for demographics. As a result, there were sex and age differences and different racial distributions between the groups. The cohorts were of insufficient size to permit sub-group comparisons. The different dosing regimens between the vaccines resulted in slightly different intervals post vaccination for sampling and could have influenced the observed immunogenicity of the various vaccines. Despite these limitations, our findings are consistent with published efficacy data for these three vaccines and provide a direct side-by-side assessment of the elicited immune responses.

Vaccination can provide protection from infection, disease, and/or death. All of these are achievable aims depending on the level and type of immunity induced by the vaccine. Moreover, protection from infection and potentially from disease also is likely to decrease transmission. As all these vaccine effects contribute to the control of the COVID-19 pandemic, a given vaccine can be effective in advancing public health goals without necessarily inducing the highest magnitude of immune response.

## Data Availability

All data are included in the manuscript.

## Acknowledgements

The authors acknowledge Saran Kupul and Lucas Schratz for assistance with sample processing, Melissa Andrasko, Spenser Kinsey, Lisa Vavro, Lalicia Roman, Jennifer Opal and Shannon Mance for assisting with subject enrollment and specimen collection, Pediatric Infectious Disease clinical research coordinators Samar Musa, Sophia Kainaroi and Isaac Cason for assistance with scheduling, and the UPMC Children’s Hospital Laboratory for assistance with subject phlebotomy

## Author Contributions

DJB: data acquisition and analysis, manuscript review and editing

EC: data acquisition, manuscript review and editing

ED: data acquisition, manuscript review and editing

JD: data acquisition and analysis, manuscript review and editing

TD: study concept and design, funding acquisition, manuscript review and editing

AH: study concept and design, funding acquisition, data acquisition, manuscript review and editing

JVW: study concept and design, funding acquisition, manuscript review and editing

MGM: study concept and design, manuscript review and editing

JFA: study concept and design, data acquisition, manuscript review and editing

WPD: study concept and design, funding acquisition, manuscript review and editing

JMM: study concept and design, IRB approval, data acquisition, study oversight, funding acquisition, manuscript review and editing

AKM: study concept and design, IRB approval, study oversight, funding acquisition, data acquisition, analysis and interpretation, manuscript writing, review and editing,

